# A novel c.515delC *HSPB8*-multisystem proteinopathy associated with inclusion body myopathy with cardiomyopathy

**DOI:** 10.1101/2024.07.17.24305631

**Authors:** Jiayan Tan, Han Duong, Margherita Milone, Liliane Gibbs, Barbara Tedesco, Angelo Poletti, Virginia Kimonis

**Affiliations:** Division of Genetics and Genomic Medicine, Department of Pediatrics; Western University of Health Sciences, College of Osteopathic Medicine of the Pacific, Pomona, CA, 91766, United States; Department of Neurology, Mayo Clinic, Rochester, Minnesota, USA; Pediatric Radiology, Department of Radiology, University of California Irvine; Laboratory of Experimental Biology, Dipartimento di Scienze Farmacologiche e Biomolecolari “Rodolfo Paoletti” (DiSFeB), Dipartimento di Eccellenza 2018–2027, Università degli Studi di Milano, 20133, Milan, Italy; Department of Neurology, Columbia University Medical Center, New York, NY, U.S.A.; Department of Neurology, University of California, Irvine, Orange CA 9286, Irvine, United States; Department of Pathology, University of California, Irvine, Orange CA 9286, Irvine, United States

**Keywords:** HSPB8, neuropathy, myopathy, cardiomyopathy, chaperone-assisted selective autophagy

## Abstract

Mutations in the *HSPB8*, or heat shock protein family B (small) member 8, is a member of the chaperone-assisted selective autophagy complex and has recently been associated with rimmed vacuolar myopathy. We report a family with a *HSPB8* rimmed vacuolar myopathy caused by a novel c.515delC variant with progressive muscle weakness, pathological findings of rimmed vacuoles of muscle biopsy, and one individual who died of cardiomyopathy. Whole exome sequencing analyses revealed a c.515delC, resulting in a translational frameshift expected to elongate the protein by an additional 49 amino acid tail. We reviewed the clinical characteristics of all reported patients (N= 26, 15 males and 11 females) in the literature, and performed a genotype/phenotype analysis. Males in the families appeared to present with an earlier age of onset, more severe muscle weakness compared to females, and a higher prevalence of other organ system complications including cardiomyopathy in two males. In conclusion, this family expands the phenotype/genotype correlations in *HSPB8*-associated myopathy with rimmed vacuoles and c.515 delC. We note that c.515 appears to be a hot spot, and the phenotype appears to be more severe in males in this disease, with an earlier onset of the myopathy.

## 1. Introduction

Chaperone-assisted selective autophagy (CASA) is a protein-mediated degradation pathway crucial for the maintenance of neuromuscular tissue [1]. HSPB8, or small heat shock protein family B member 8 complexes with co-chaperone BAG3, and together they associate with the complex formed by HSC70, a selected member of the heat shock protein family A (HSPA), E3 ubiquitin ligase STIP1 homology, and U-Box containing protein 1 (STUB1, also known as C terminus of HSC70-Interacting Protein, CHIP). This heteromeric complex is known as the CASA complex [2–5]. HSPB8 aids in recognizing misfolded proteins, guiding the proteins to refold via HSPA or degradation via STUB1-mediated ubiquitination [4]. Mutations in genes that disrupt the CASA pathway are associated with central nervous system, and neuromuscular disorders [6–17]. Failure in protein degradation result in BAG3-mediated shunting to aggresomes ^1^[18]. *HSPB8* mutations are associated with Charcot-Marie-Tooth type 2 L (CMT2L), distal hereditary motor neuropathy type II (dHMNII), and recently myopathy with rimmed vacuolar and myofibrillar pathology [19–25]. Frameshifts in the *HSPB8* gene, resulting in elongated protein sequence, have been demonstrated to form insoluble protein aggregates, disrupting the CASA complex formation and function [1]. Here we present a family with an *HSPB8* frameshift mutation c.515delC manifesting as rimmed vacuolar myopathy, and compared their findings with all previous reports of *HSPB8* myofibrillar myopathy to expand the clinical phenotype of this rare disease [20–28].

## 2. Methods

This study was approved by the University of California Irvine Institutional Review Board (IRB 2007-5832). We obtained written consent for the studies. Whole exome sequencing was performed on the proband. The proband and his family’s clinical histories were reviewed. A literature review of all existing *HSPB8* variant cases were reviewed as comparison.

## 3. Results

The patient is a male in his thirties. The patient reported difficulty running in childhood. In high school, the proband was diagnosed with scoliosis and flat feet. In his twenties his chest x-ray showed scoliosis and MRI revealed severe thoracic paraspinal muscle atrophy (Fig. 1, 2). In his late twenties the patient reported foot and ankle pain, progressive muscle weakness, and increased difficulty going up the stairs, which prompted him to see a neurologist.

**Figure 1:**
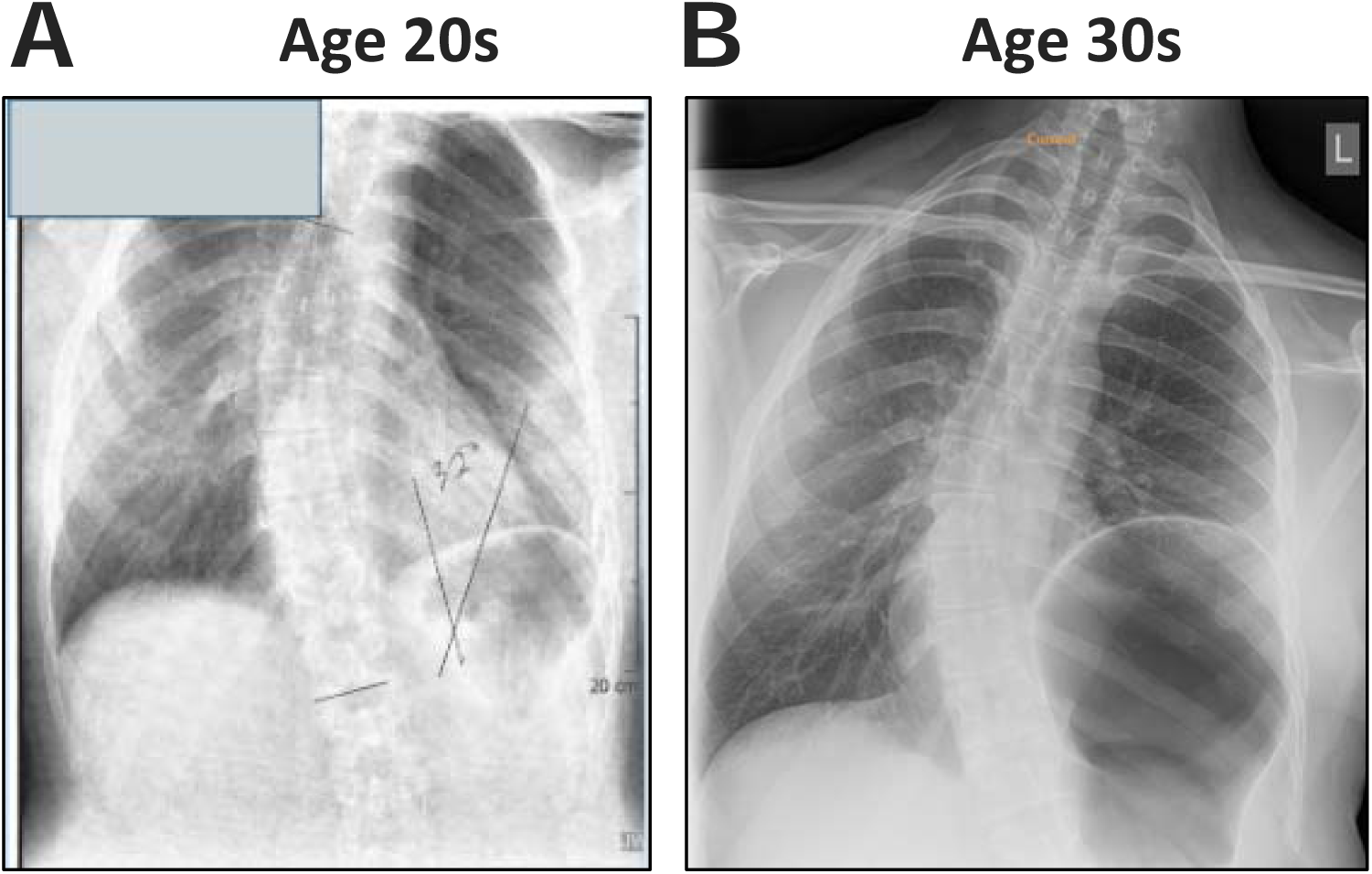
X-ray of the proband’s thoracolumbar spine. (A and B) X-ray of the proband’s spine in his twenties and thirties shows increase in scoliotic curvature and left hemidiaphragm elevation.

Using the Medical Research Council (MRC) grading scale, the patient, aged 31-35 y., demonstrates predominantly normal muscle strength, graded as 5/5, with exceptions including slight weakness graded as 4+/5 in right elbow extension, moderate weakness graded as 3/5 in hip flexion, extension, and adduction, and significant weakness graded as 2/5 in right dorsiflexion, and 3+/5 on the left. Reflexes including the biceps, triceps, brachioradialis, knee, and ankles were normal. The sensation exam was unremarkable. He also experienced asymmetric abdominal muscle weakness, rhomboid area atrophy, and a positive Beevor’s sign [29]. Needle electromyography revealed rapid recruitment of short, low amplitude motor unit potentials with fibrillation potentials in proximal, distal and axial muscles. Sensory nerve conductions were normal.

On physical exam, there was severe atrophy in the paraspinal, rhomboid, abdominal, psoas, gluteal, and leg muscles, with relative sparing of the medial and lateral head of the gastrocnemius. There was a progressive bilateral scapular winging with increasing proximal leg weakness. The patient has a moderate S-shaped scoliosis of thoracolumbar spine (thoracic dextroscoliosis and lumbar levoscoliosis), increased lumbar lordosis, and an anterior pelvic tilt (Fig. 1).

In his twenties the patient started to experience mild dyspnea walking up the stairs. His pulmonary function tests (PFT) showed forced vital capacity (FVC) of 69% (normal >80%), forced expiratory volume (FEV1) of 68% (normal >80%), and total lung capacity (TLC) of 72% (normal >80%), revealing a restrictive lung defect [30]. At age 36-40, his PFTs were worsening based on FVC of 45%, FEV1 of 47%, and TLC of 45%. His chest x-ray showed elevation of the left diaphragm from eventration thinning, indicating hemidiaphragm paralysis (Fig. 1). Bilevel positive airway pressure (BiPAP) via face mask was recommended for his respiratory insufficiency.

Cardiovascular assessment, echocardiogram (ECG), and 24-hour ECG Holter monitoring revealed no abnormalities. Cardiac magnetic resonance imaging (MRI) indicated normal function, anatomy, and a non-clinically relevant pericardial effusion with normal pericardial thickness.

The proband in his early 30s exhibited consistently elevated total triglycerides (196 mg/dL, normal < 150 mg/dL) and low high-density lipoprotein (HDL) cholesterol (24 mg/dL, normal 40-60 mg/dL) [31]. Laboratory findings showed normal comprehensive metabolic profile, alkaline phosphatase, and liver function tests. Muscle enzyme analysis revealed elevated total creatine kinase (237 U/L, normal < 170 U/L) and creatine kinase-myoglobin binding (CK-MB) (12.7 ng/mL, normal < 5.0 μg/L) [32].

His past medical history includes flat feet, scoliosis, dyspnea with stairs, hypertension, hypercholesterolemia, sleep apnea, pericardial effusion, non-alcoholic fatty liver disease (NAFLD), acquired pyloric stenosis, and elevated left hemidiaphragm and eventration.

### 3.1 Imaging Studies

At age 21-25 y, the patient’s lumbar MRI showed severe paraspinal atrophy with fatty infiltration (Fig. 2). Additionally, the thigh MRIs showed fat infiltration of the majority of the lower extremity muscles with marked and symmetric atrophy of bilateral thigh muscles but mild relative sparing of rectus femoris and vastus lateralis (Fig. 2). The calf MRIs showed moderate muscle atrophy with relative sparing of medial and lateral heads of gastrocnemius (Fig. 2).

**Figure 2:**
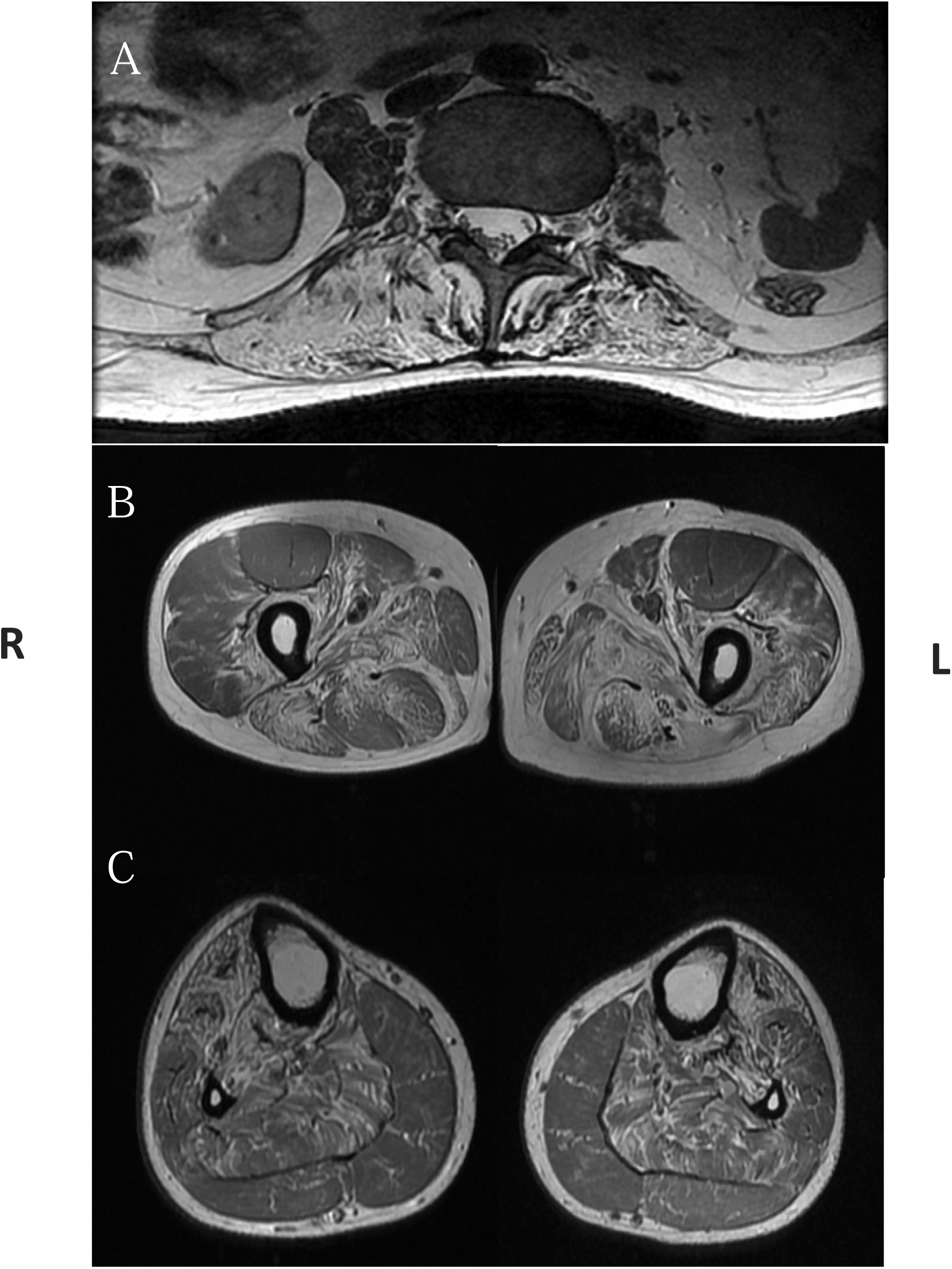
Lumbar and Leg MRI in his thirties showing marked muscle atrophy and fatty infiltration in paraspinal and hamstring muscle. (A) Lumbar MRI showed paraspinal, psoas, and gluteal muscle atrophy, (B) Right and left thigh showed marked hamstring atrophy with relative sparing of the rectus femoris, vastus lateralis, gracilis, and sartorius muscles, (C) Right and left calf MRI showed fatty infiltration and atrophy with relative sparing of gastrocnemius and peroneus muscles.

Cardiac MRI incidentally detected left lower lobe atelectasis and elevation of the left hemidiaphragm. Abdomen MRI showed hepatomegaly, grade 1 hepatic steatosis, and some evidence of fibrosis.

### 3.2 Muscle Histology

The biopsy of the gracilis showed a marked increase in internalized nuclei, a few fibers subdividing by splitting, a few regenerating and rare necrotic fibers. Several fibers harbored vacuoles rimmed by membranous material while other fibers contained small eosinophilic inclusions. In trichrome stained sections, several fibers showed hyaline inclusions. Endomysial connective tissue was mildly to moderately increased in different fascicles. There was a minimal collection of inflammatory cells at a perivascular site. In ATPase reacted sections, there was no fiber type grouping, and the few atrophic fibers were of either histochemical type. No inclusions were noted in Congo red stained sections, but this stain was performed on paraffin-embedded tissue. Muscle biopsy of the left quadriceps revealed increased internalized nuclei and exceedingly rare, rimmed vacuoles. The left obturator nerve biopsy showed normal density myelinated nerve fibers with mild dropout.

### 3.3 Family History

The patient has a strong family history of autosomal dominant muscle weakness in his parent, grandparent, and relative (Fig. 3). The parent developed mild muscle weakness during her 40s-50s and struggled with walking in her 60s. A muscle biopsy retrieved during her hip replacement revealed myofibrillar disruption. The grandparent had proximal limb and distal leg weaknesses and died in her late 80s. The proband’s relative was diagnosed with ‘muscular dystrophy’, had to use a cane in his late 20s, used a wheelchair in his 30s, and was unable to lift arms in his 40-50s. The proband’s relative had a muscle biopsy done that revealed rimmed vacuoles. His echocardiogram showed left ventricular hypertrophy, reduced ejection fraction, and mild mitral regurgitation and he died of ‘cardiomyopathy’ in his early fifties

**Figure 3:**
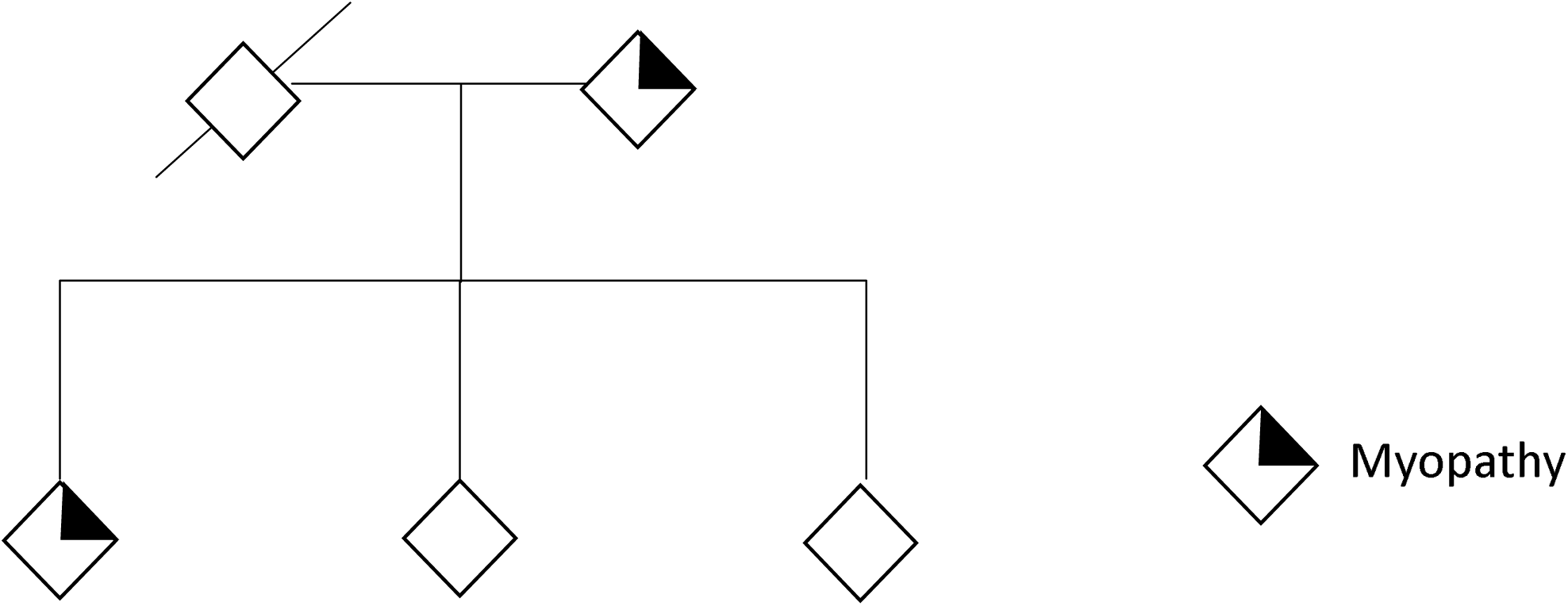
Family pedigree showing autosomal dominant inheritance. The proband and relative demonstrate a history of muscle weakness. Asterisk indicates genetic testing result of *HSPB8* c.515delC. (Please contact Dr. Virginia Kimonis for access to details of the pedigree.)

### 3.4 Genetic Testing

The patient underwent an Limb Girdle Muscular Dystrophy (LGMD) panel test, which revealed 6 variants of unknown significance (VUS) in DYSF (c.4198C>G, p.Pro1400Ala), PLEC (c.8263A>G, p.Ile2755Val), TCAP (c.37_39:3 bp del of GAG, Codon:13), TRIM32 (c.127A>G, p.Ile43Val), TTN (c.104251G>C, p.Ala34751Pro), and ANO5 (c.720G>T).

Dysferlinopathy was ruled out through normal dysferlin expression levels, and FSHD1 was excluded by a negative D4Z4 repeat contraction on Chr 4q35. Further investigation via whole-exome trio sequencing identified a frameshift mutation (c.515delC; p.Pro172Leufs*75) in the *HSPB8* gene, confirmed in a Clinical Laboratory Improvement Amendments of 1988 (CLIA) - certified laboratory (Fig. 4).

**Figure 4:**
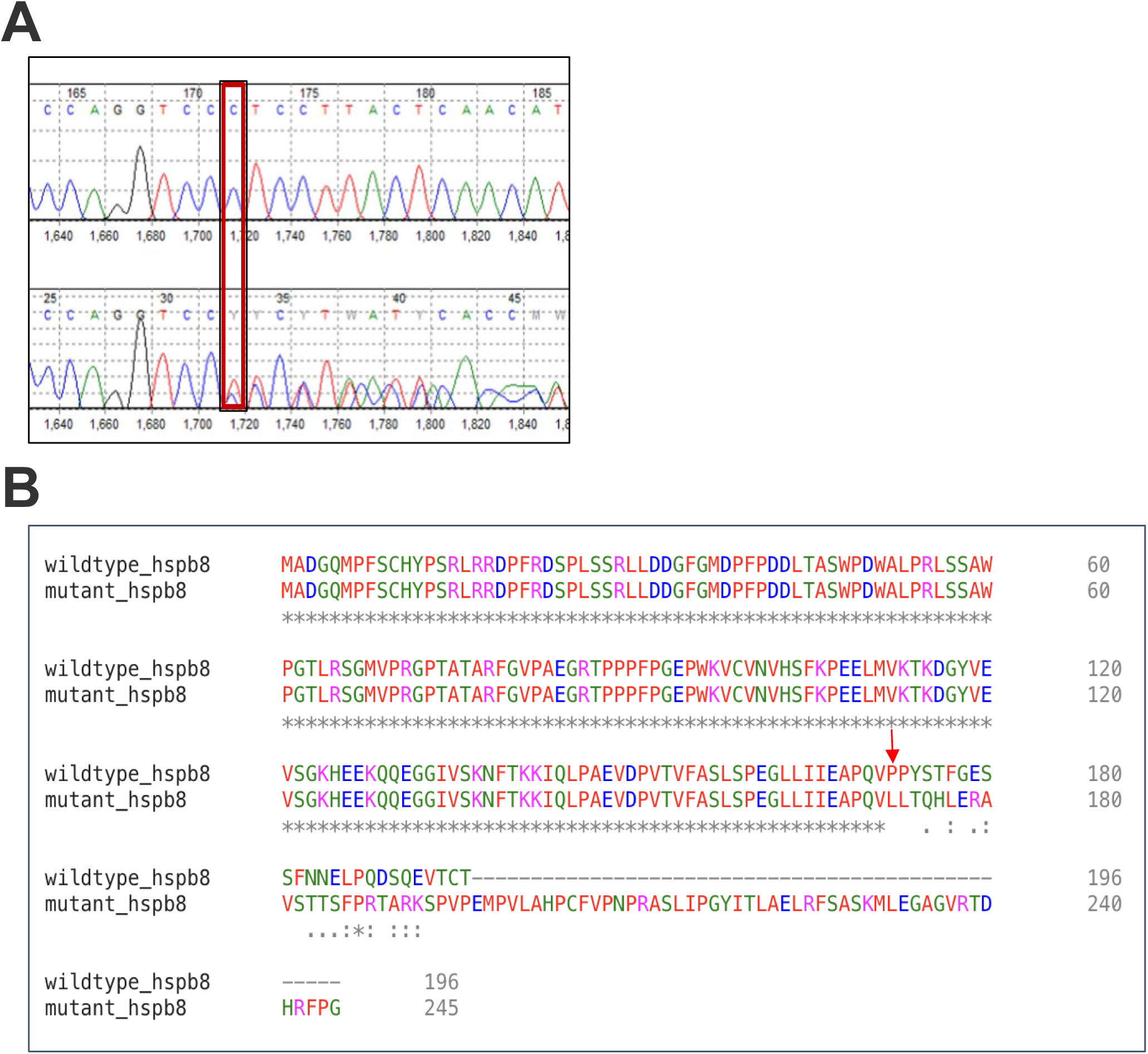
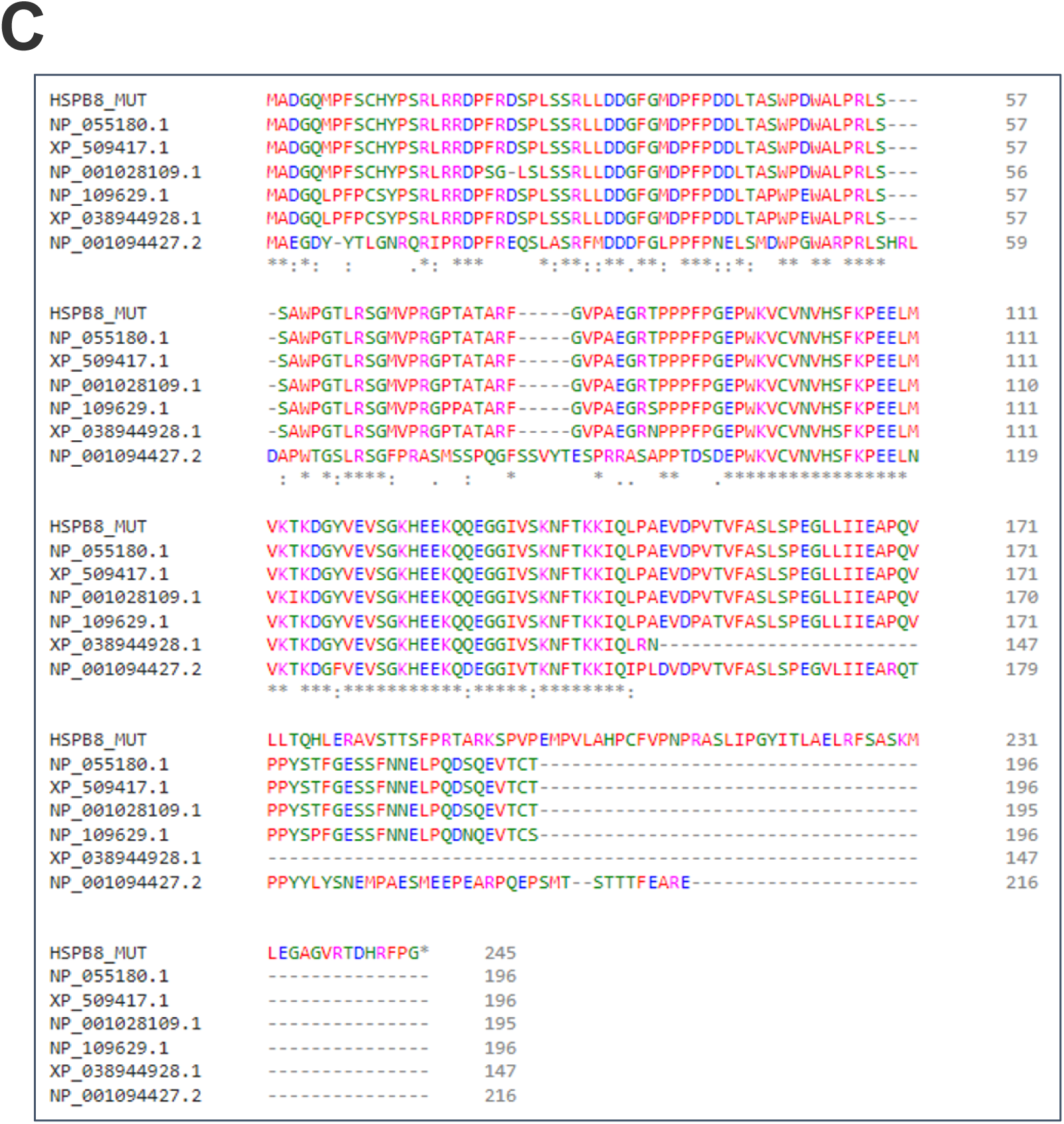
Chromatogram and alignment of the familial pathogenic *HSPB8* c.515delC variant. (A) Chromatogram from the proband revealing the frameshift mutation c.515delC; p.Pro172Leufs*75 (red rectangles), which was similarly identified in the patient. (B) Alignment of the mutated protein and the normal HSPB8 protein in human (NP_055180.1). The mutated gene codifies a protein with a frameshift of 25 amino acids and an additional extension of 49 amino acids. (C) Alignment of the mutated protein (HSPB8_MUT) and the normal HSPB8 protein in human (NP_055180.1), chimpanzee (XP_509417.1), rhesus macaque (NP_001028109.1), mouse (NP_109629.1), rat (XP_038944828.1) and zebrafish (NP_001094427.2). The mutated gene codifies a protein with a frameshift of 25 amino acids and an additional extension of 49 amino acids. The affected residues are highly conserved between the analyzed species.

The *HSPB8* gene deletion of C at position 515, located in coding exon 3, the last coding exon, causes a translational frameshift with a predicted alternate stop codon (p.Pro172Leufs*75). This deletion results in a frameshift at amino acid 172/197 and is expected to disrupt the last 25 amino acids of HSPB8 protein and extend the protein by an additional 49 amino acids. The affected residues are conserved among various species, including chimpanzee, rhesus macaque, mouse, rat, and zebrafish (Fig. 4). Given the deleterious nature of the c.515delC (p.Pro172Leufs*75) variant identified in this patient, and its absence in population databases (GnomAD and ExAC), it is reported as likely pathogenic. The calculated severity score is 1.0 on a scale of 0.0 – 1.0.

### 3.5 Literature review of all existing HSPB8 myopathy

In this study, we systematically examined reported clinical findings and *HSPB8* variants in individuals, explored phenotypic differences between *HSPB8* variants, and the influence of sex in six families and five sporadic cases reported to have *HSPB8* variants (Table. 1). Of the 26 cases, 15 (58%) were male and 11 (42%) were female. The median age of onset of the myopathy was 35 y. in males and 38.5 y. in females, with a range of 19-56 y. and 20-46 y., respectively.

**Table 1:**
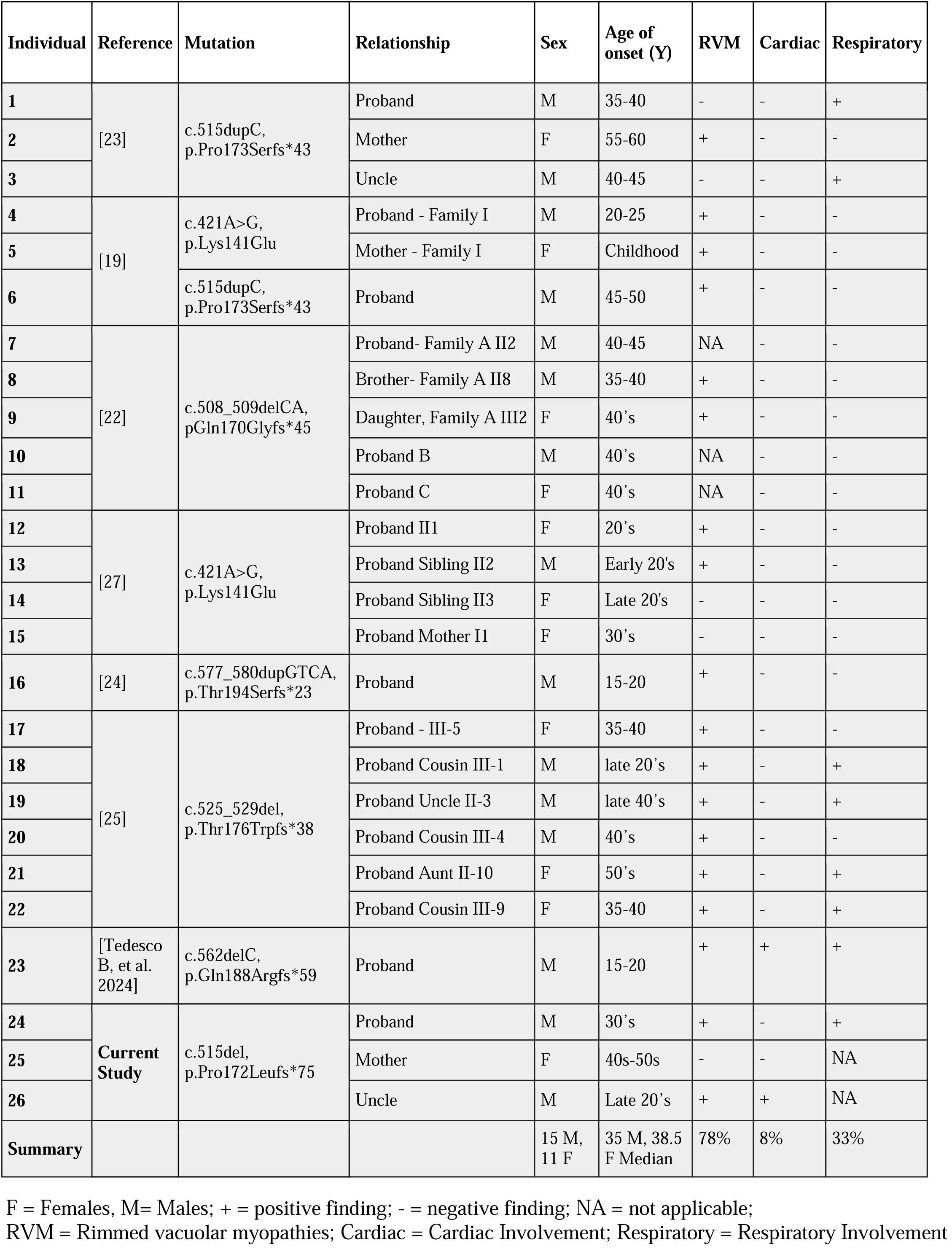
Comparisons of all reported HSPB8 cases with current study.

Eighteen out of twenty-four (78%) individuals had rimmed vacuolar myopathies (RVMs), of which 10 (59%) were male and seven (41%) were female. Of the eight out of twenty-four (33%) individuals reported to have respiratory insufficiency/failure, six (75%) were male and two (25%) were female. Two individuals were reported to have cardiomyopathy complications, and both were males (100%).

We note that the c.515 codon appears to be a hot spot. A variant in the same codon 515dupC, p.P173Sfs*43 had been shown to be associated with a progressive distal myopathy and neuropathy with muscle tissue showed increased internal nuclei, numerous rimmed vacuolar fibers, splitting, cytoplasmic bodies, and moth-eaten fibers in two families [19 23]. Frameshift variants have been reported two amino acids upstream [28] and at the adjacent amino acid downstream in patients with distal motor neuropathy[19 23].

## 4. Discussion

We report a family of Ashkenazi Jewish descent with hereditary rimmed vacuolar myopathy associated with a novel variant in the *HSPB8* gene [1]. The proband has proximal, distal, and axial muscle weakness. Other phenotypes include restricted lung defect, left elevated hemidiaphragm, and severe scoliosis. His parent has a similar but milder phenotype and his parent’s sibling passed from cardiomyopathy. The deletion of C at position 515 of the *HSPB8* gene causes a translational frameshift with a predicted alternate stop codon (p.Pro172Leufs*75), predicted to disrupt the last 25 amino acids of HSPB8 protein and extend the protein an additional 49 amino acid tail. Most reported *HSPB8* mutations occur within the α-crystallin domain that manifest chaperone activity [23].

Here, we report individuals in the same family sharing a c.515delC, p.Pro172Leufs*75 variant of *HSPB8*. We also analyzed and compared the clinical features to seven individuals within two other families who have a mutation in the *HSPB8* hotspot c. 515 position (c.515dupC, p.Pro173Serfs*43). Al-Tahan et al. (2019) reported three individuals and Ghaoui et al. (2019) reported one sporadic case of the same variant, both presenting with predominantly distal neuromyopathy (quadricep > hamstring) characterized by myofibrillar aggregates, rimmed vacuoles, and neurogenic features on biopsy and electromyographic studies [19, 23], unlike our family who feature predominantly proximal weakness. The age of onset is also earlier for the c.515delC group, with affected males experiencing onset in their late 20s to 30s, compared to later age of onset ranging from 35 to 46 y. in the dupC group. The females in the two groups manifested in the 40s in the delC group versus 50s. in the dupC group.

### 4.1 Gender-associated myopathy

Males with the *HSPB8* variants had an earlier onset of myopathy (median age onset in the mid thirties) versus females (median age high thirties.) with more severe complications. Males also had a higher incidence of RVMs, respiratory insufficiency, and cardiomyopathy, compared to females. The mechanism underlying the gender discrepancy in clinical presentation is still under investigation. One potential mechanism could be related to the protective influence of estrogens, potentially mediated by HSPB8 and other CASA complex molecules. Indeed, estrogens are known to be HSPB8 inducers, the enhancement is prevented by stimulation of the Estrogen Receptor (ER) antagonist [33–36]. Other female hormones like progestins also function as HSPB8 inducers [33 34 37]. Also reported was the potential role of ER (/3 form) in regulating the expression of autophagy markers, light chain 3 (LC3-I/LC3-II), and sequestosome-1 (SQSTM1/p62), which are key autophagy molecules in the CASA complex [38]. There is also an association between estrogen and the preservation of muscular structure and function in mice models treated with ER agonist [39]. Moreover, post-menopausal women have an increased risk of musculoskeletal injury and muscle wasting [40]. On the other hand, one may consider that muscle in males is under continuous stimulation of androgens acting as anabolic steroids via the androgen receptor (AR) [41]; it is unclear whether this stimulation may make male skeletal muscle more sensitive than female skeletal muscle to deleterious activities of the mutated *HSPB8*. In response to these findings, interventions such as hormone replacement therapy have been employed to counteract the adverse effects of menopause, reduce muscle and bone loss while restoring muscle protein balance [42 43].

### 4.2 HSPB8 & Cardiomyopathy

HSPB8 expression is highly enriched in cardiac, skeletal, and smooth muscle tissues [44]. Studies suggested its role in maintaining proteostasis to prevent damage to cardiomyocytes [45]. An experimental study involving a mutated *HSPB8* gene in mice showed distal hereditary motor neuropathy, mild ventricular hypertrophy, and apical cardiac fibrosis [46]. Studies have shown disruption of the CASA complex in patient and animal models causes myopathy and cardiomyopathy [10 19 46]. BAG3 variants identified in humans are associated with cardiomyopathy [47 48]. Additionally in animal studies, *Bag3*−/− mice were reported to have abnormalities in skeletal and cardiac tissue [10 19 49]. Cardiomyopathy was reported in two individuals with pathogenic *HSPB8* variant, however, additional studies in a larger cohort is needed to establish this association, or by excluding other genetic etiologies.

### 4.3 HSPB8 & Multisystem Proteinopathy

In a recent report, *HSPB8* was listed as a potential multisystem proteinopathy gene, with association of motor neuropathy, sensory neuropathy, and rimmed vacuoles [50]. Other MSP-causative genes include *VCP*, *HNRNPA2B1*, *HNRNPA1*, *SQSTM1, MATR3*, *TIA1*, and *TFG* [33]. Interestingly, myopathy with rimmed vacuoles is reported as the most common initial presentation for multisystem involvement of proteinopathies [50]. These disorders are also associated with neurodegenerative conditions such as amyotrophic lateral sclerosis, frontotemporal dementia, Paget disease of bone, restrictive respiratory defect, and cardiomyopathy (diastolic dysfunction and increased concentric left ventricular wall thickness). Additionally TAR DNA binding protein-43 (TDP-43) pathology is common in these disorders including RVM associated with *HSPB8* variants. TDP-43 is known to colocalize with other proteins like p62, contributing to the formation of myofibrillar aggregates and possibly rimmed vacuoles [19 23 27 50].

## 5. Conclusion

In conclusion, this paper explores the clinical and molecular aspects of a novel frameshift mutation (c.515delC; p.Pro172Leufs*75) in the *HSPB8* gene, potentially contributing to a multisystemic manifestation of muscular disorders, including respiratory insufficiency and possibly cardiomyopathy. The *HSPB8* c.515 variant appears to be a mutation hot spot since two other families have been reported with c.515 dup variants at this locus. From analyzing 26 cases of individuals with *HSPB8* variants and myopathy, we see a later onset in females than males, with age of onset 38.5 y. versus 35 y., and milder symptoms, suggesting that gender plays a role in the differences in phenotypic expression of *HSPB8*-related myopathy.

This family not only contributes a detailed clinical characterization of a unique *HSPB8* frameshift mutation case but also underscores the importance of considering *HSPB8* in the differential diagnosis of neuromuscular disorders with multisystemic involvement. The findings pave the way for future research into the common molecular mechanisms underlying *HSPB8* and related multisystem proteinopathies.

## Data Availability

All data produced in the present work are contained in the manuscript

## 1 Abbreviations

AA: Amino Acids
AR: Androgen Receptor
BiPAP: Bilevel Positive Airway Pressure
CASA: Chaperone-assisted selective autophagy
CK-MB: Creatine Kinase-Myoglobin Binding
CLIA: Clinical Laboratory Improvement Amendments of 1988
CMT2L: Charcot-Marie-Tooth type 2 L
dHMNII: Distal Hereditary Motor Neuropathy Type II
ER: Estrogen Receptor
FEV1: Forced Expiratory Volume
FVC: Forced Vital Capacity
HDL: High Density Lipoprotein
HSPB8: Heat Shock Protein Family B (small) Member 8
HSPA: Heat Shock Protein Family A
LC3-I/LC3-II: Light Chain 3
RVM: Rimmed Vacuolar Myopathy
MRI: Magnetic Resonance Imaging
MRC: Medical Research Council
MSP: Multisystem Proteinopathy
NAFLD: Non-Alcoholic Fatty Liver Disease
PFT: Pulmonary Function Test
SQSTM1/p62: Sequestosome-1
TDP-43: TAR DNA Binding Protein-43
TLC: Total Lung Capacity
VUS: Variants of Unknown Significance

## Acknowledgment

We thank the proband and his family members for permitting us to report their medical history and imaging. We also thank Yousef Award for assisting with data analysis. We thank Drs. Michio Hirano, and Matthew Harms for their assistance with data acquisition.

## References

1. Tedesco B, Vendredy L, Adriaenssens E, et al. HSPB8 frameshift mutant aggregates weaken chaperone-assisted selective autophagy in neuromyopathies. Autophagy 2023;19(8):2217–39 doi: 10.1080/15548627.2023.2179780 [published Online First: 20230228].

2. Arndt V, Dick N, Tawo R, et al. Chaperone-assisted selective autophagy is essential for muscle maintenance. Curr Biol 2010;20(2):143–8 doi: 10.1016/j.cub.2009.11.022 [published Online First: 20100107].

3. Carra S, Seguin SJ, Lambert H, Landry J. HspB8 chaperone activity toward poly(Q)-containing proteins depends on its association with Bag3, a stimulator of macroautophagy. J Biol Chem 2008;283(3):1437–44 doi: 10.1074/jbc.M706304200 [published Online First: 20071115].

4. Rauch JN, Tse E, Freilich R, et al. BAG3 Is a Modular, Scaffolding Protein that physically Links Heat Shock Protein 70 (Hsp70) to the Small Heat Shock Proteins. J Mol Biol 2017;429(1):128–41 doi: 10.1016/j.jmb.2016.11.013 [published Online First: 20161121].

5. Ballinger CA, Connell P, Wu Y, et al. Identification of CHIP, a novel tetratricopeptide repeat-containing protein that interacts with heat shock proteins and negatively regulates chaperone functions. Mol Cell Biol 1999;19(6):4535–45 doi: 10.1128/MCB.19.6.4535.

6. Jaffer F, Murphy SM, Scoto M, et al. BAG3 mutations: another cause of giant axonal neuropathy. J Peripher Nerv Syst 2012;17(2):210–6 doi: 10.1111/j.1529-8027.2012.00409.x.

7. Kostera-Pruszczyk A, Suszek M, Ploski R, et al. BAG3-related myopathy, polyneuropathy and cardiomyopathy with long QT syndrome. J Muscle Res Cell Motil 2015;36(6):423–32 doi: 10.1007/s10974-015-9431-3 [published Online First: 20151106].

8. Lee HC, Cherk SW, Chan SK, et al. BAG3-related myofibrillar myopathy in a Chinese family. Clin Genet 2012;81(4):394–8 doi: 10.1111/j.1399-0004.2011.01659.x [published Online First: 20110404].

9. Meister-Broekema M, Freilich R, Jagadeesan C, et al. Myopathy associated BAG3 mutations lead to protein aggregation by stalling Hsp70 networks. Nat Commun 2018;9(1):5342 doi: 10.1038/s41467-018-07718-5 [published Online First: 20181217].

10. Selcen D, Muntoni F, Burton BK, et al. Mutation in BAG3 causes severe dominant childhood muscular dystrophy. Ann Neurol 2009;65(1):83–9 doi: 10.1002/ana.21553.

11. Semmler AL, Sacconi S, Bach JE, et al. Unusual multisystemic involvement and a novel BAG3 mutation revealed by NGS screening in a large cohort of myofibrillar myopathies. Orphanet J Rare Dis 2014;9:121 doi: 10.1186/s13023-014-0121-9 [published Online First: 20140801].

12. Shy M, Rebelo AP, Feely SM, et al. Mutations in BAG3 cause adult-onset Charcot-Marie-Tooth disease. J Neurol Neurosurg Psychiatry 2018;89(3):313–15 doi: 10.1136/jnnp-2017-315929 [published Online First: 20170728].

13. Villard E, Perret C, Gary F, et al. A genome-wide association study identifies two loci associated with heart failure due to dilated cardiomyopathy. Eur Heart J 2011;32(9):1065–76 doi: 10.1093/eurheartj/ehr105 [published Online First: 20110401].

14. Cordoba M, Rodriguez-Quiroga S, Gatto EM, Alurralde A, Kauffman MA. Ataxia plus myoclonus in a 23-year-old patient due to STUB1 mutations. Neurology 2014;83(3):287–8 doi: 10.1212/WNL.0000000000000600 [published Online First: 20140613].

15. Heimdal K, Sanchez-Guixe M, Aukrust I, et al. STUB1 mutations in autosomal recessive ataxias - evidence for mutation-specific clinical heterogeneity. Orphanet J Rare Dis 2014;9:146 doi: 10.1186/s13023-014-0146-0 [published Online First: 20140926].

16. Shi Y, Wang J, Li JD, et al. Identification of CHIP as a novel causative gene for autosomal recessive cerebellar ataxia. PLoS One 2013;8(12):e81884 doi: 10.1371/journal.pone.0081884 [published Online First: 20131202].

17. Mengel D, Traschutz A, Reich S, et al. A de novo STUB1 variant associated with an early adult-onset multisystemic ataxia phenotype. J Neurol 2021;268(10):3845–51 doi: 10.1007/s00415-021-10524-7 [published Online First: 20210403].

18. Gamerdinger M, Kaya AM, Wolfrum U, Clement AM, Behl C. BAG3 mediates chaperone-based aggresome-targeting and selective autophagy of misfolded proteins. EMBO Rep 2011;12(2):149–56 doi: 10.1038/embor.2010.203 [published Online First: 20110121].

19. Ghaoui R, Palmio J, Brewer J, et al. Mutations in HSPB8 causing a new phenotype of distal myopathy and motor neuropathy. Neurology 2016;86(4):391–8 doi: 10.1212/WNL.0000000000002324 [published Online First: 20151230].

20. Irobi J, Van Impe K, Seeman P, et al. Hot-spot residue in small heat-shock protein 22 causes distal motor neuropathy. Nat Genet 2004;36(6):597–601 doi: 10.1038/ng1328 [published Online First: 20040502].

21. Nakhro K, Park JM, Kim YJ, et al. A novel Lys141Thr mutation in small heat shock protein 22 (HSPB8) gene in Charcot-Marie-Tooth disease type 2L. Neuromuscul Disord 2013;23(8):656–63 doi: 10.1016/j.nmd.2013.05.009 [published Online First: 20130621].

22. Echaniz-Laguna A, Geuens T, Petiot P, et al. Axonal Neuropathies due to Mutations in Small Heat Shock Proteins: Clinical, Genetic, and Functional Insights into Novel Mutations. Hum Mutat 2017;38(5):556–68 doi: 10.1002/humu.23189 [published Online First: 20170225].

23. Al-Tahan S, Weiss L, Yu H, et al. New family with HSPB8-associated autosomal dominant rimmed vacuolar myopathy. Neurol Genet 2019;5(4):e349 doi: 10.1212/NXG.0000000000000349 [published Online First: 20190710].

24. Nicolau S, Liewluck T, Elliott JL, Engel AG, Milone M. A novel heterozygous mutation in the C-terminal region of HSPB8 leads to limb-girdle rimmed vacuolar myopathy. Neuromuscul Disord 2020;30(3):236–40 doi: 10.1016/j.nmd.2020.02.005 [published Online First: 20200212].

25. Inoue-Shibui A, Niihori T, Kobayashi M, et al. A novel deletion in the C-terminal region of HSPB8 in a family with rimmed vacuolar myopathy. J Hum Genet 2021;66(10):965–72 doi: 10.1038/s10038-021-00916-y [published Online First: 20210320].

26. Ghaoui R, Palmio J, Brewer J, et al. Mutations in HSPB8 causing a new phenotype of distal myopathy and motor neuropathy. Neurology 2016;86(4):391–8 doi: 10.1212/WNL.0000000000002324.

27. Cortese A, Laura M, Casali C, et al. Altered TDP-43-dependent splicing in HSPB8-related distal hereditary motor neuropathy and myofibrillar myopathy. Eur J Neurol 2018;25(1):154–63 doi: 10.1111/ene.13478 [published Online First: 20171202].

28. Echaniz-Laguna A, Lornage X, Lannes B, et al. HSPB8 haploinsufficiency causes dominant adult-onset axial and distal myopathy. Acta Neuropathol 2017;134(1):163–65 doi: 10.1007/s00401-017-1724-8 [published Online First: 20170513].

29. Althagafi A, Nadi M. Beevor Sign. StatPearls. Treasure Island (FL), 2024.

30. Ponce MC, Sankari A, Sharma S. Pulmonary Function Tests. StatPearls. Treasure Island (FL), 2024.

31. Lee Y, Siddiqui WJ. Cholesterol Levels. StatPearls. Treasure Island (FL), 2024.

32. Cabaniss CD. Creatine Kinase. In: Walker HK, Hall WD, Hurst JW, eds. Clinical Methods: The History, Physical, and Laboratory Examinations. 3rd ed. Boston, 1990.

33. Sun X, Fontaine JM, Bartl I, Behnam B, Welsh MJ, Benndorf R. Induction of Hsp22 (HspB8) by estrogen and the metalloestrogen cadmium in estrogen receptor-positive breast cancer cells. Cell Stress Chaperones 2007;12(4):307–19 doi: 10.1379/csc-276.1.

34. Cristofani R, Piccolella M, Crippa V, et al. The Role of HSPB8, a Component of the Chaperone-Assisted Selective Autophagy Machinery, in Cancer. Cells 2021;10(2) doi: 10.3390/cells10020335 [published Online First: 20210205].

35. Piccolella M, Cristofani R, Tedesco B, et al. Retinoic Acid Downregulates HSPB8 Gene Expression in Human Breast Cancer Cells MCF-7. Front Oncol 2021;11:652085 doi: 10.3389/fonc.2021.652085 [published Online First: 20210531].

36. Piccolella M, Crippa V, Cristofani R, et al. The small heat shock protein B8 (HSPB8) modulates proliferation and migration of breast cancer cells. Oncotarget 2017;8(6):10400–15 doi: 10.18632/oncotarget.14422.

37. Dressing GE, Knutson TP, Schiewer MJ, et al. Progesterone receptor-cyclin D1 complexes induce cell cycle-dependent transcriptional programs in breast cancer cells. Mol Endocrinol 2014;28(4):442–57 doi: 10.1210/me.2013-1196 [published Online First: 20140225].

38. Yang ZM, Yang MF, Yu W, Tao HM. Molecular mechanisms of estrogen receptor beta-induced apoptosis and autophagy in tumors: implication for treating osteosarcoma. J Int Med Res 2019;47(10):4644–55 doi: 10.1177/0300060519871373 [published Online First: 20190917].

39. Momb BA, Szabo GK, Mogus JP, Chipkin SR, Vandenberg LN, Miller MS. Skeletal Muscle Function Is Altered in Male Mice on Low-Dose Androgen Receptor Antagonist or Estrogen Receptor Agonist. Endocrinology 2023;164(10) doi: 10.1210/endocr/bqad132.

40. Enns DL, Tiidus PM. The influence of estrogen on skeletal muscle: sex matters. Sports Med 2010;40(1):41–58 doi: 10.2165/11319760-000000000-00000.

41. Hosoi T, Yakabe M, Hashimoto S, Akishita M, Ogawa S. The roles of sex hormones in the pathophysiology of age-related sarcopenia and frailty. Reprod Med Biol 2024;23(1):e12569 doi: 10.1002/rmb2.12569 [published Online First: 20240311].

42. Hansen M, Skovgaard D, Reitelseder S, Holm L, Langbjerg H, Kjaer M. Effects of estrogen replacement and lower androgen status on skeletal muscle collagen and myofibrillar protein synthesis in postmenopausal women. J Gerontol A Biol Sci Med Sci 2012;67(10):1005–13 doi: 10.1093/gerona/gls007 [published Online First: 20120301].

43. Smith GI, Yoshino J, Reeds DN, et al. Testosterone and progesterone, but not estradiol, stimulate muscle protein synthesis in postmenopausal women. J Clin Endocrinol Metab 2014;99(1):256–65 doi: 10.1210/jc.2013-2835 [published Online First: 20131220].

44. Fang X, Bogomolovas J, Trexler C, Chen J. The BAG3-dependent and -independent roles of cardiac small heat shock proteins. JCI Insight 2019;4(4) doi: 10.1172/jci.insight.126464 [published Online First: 20190221].

45. Hu X, Van Marion DMS, Wiersma M, Zhang D, Brundel B. The protective role of small heat shock proteins in cardiac diseases: key role in atrial fibrillation. Cell Stress Chaperones 2017;22(4):665–74 doi: 10.1007/s12192-017-0799-4 [published Online First: 20170508].

46. Sanbe A, Marunouchi T, Abe T, et al. Phenotype of cardiomyopathy in cardiac-specific heat shock protein B8 K141N transgenic mouse. J Biol Chem 2013;288(13):8910–21 doi: 10.1074/jbc.M112.368324 [published Online First: 20130206].

47. Qu HQ, Feldman AM, Hakonarson H. Genetics of BAG3: A Paradigm for Developing Precision Therapies for Dilated Cardiomyopathies. J Am Heart Assoc 2022;11(23):e027373 doi: 10.1161/JAHA.122.027373 [published Online First: 20221116].

48. Dominguez F, Cuenca S, Bilinska Z, et al. Dilated Cardiomyopathy Due to BLC2-Associated Athanogene 3 (BAG3) Mutations. J Am Coll Cardiol 2018;72(20):2471–81 doi: 10.1016/j.jacc.2018.08.2181.

49. Homma S, Iwasaki M, Shelton GD, Engvall E, Reed JC, Takayama S. BAG3 deficiency results in fulminant myopathy and early lethality. Am J Pathol 2006;169(3):761–73 doi: 10.2353/ajpath.2006.060250.

50. Chompoopong P, Oskarsson B, Madigan NN, et al. Multisystem proteinopathies (MSPs) and MSP-like disorders: Clinical-pathological-molecular spectrum. Ann Clin Transl Neurol 2023;10(4):632–43 doi: 10.1002/acn3.51751 [published Online First: 20230301].

